# Konjac mannan oligosaccharides as a sustainer of fasting-associated gut microbiome signature after discontinuation of intermittent fasting in overweight individuals: A protocol for an open-label randomized clinical trial

**DOI:** 10.1101/2025.05.30.25328655

**Authors:** Junhong Su, Guorong Ma, Xianghua Tang, Zhongren Ma, Zhenrong Xie, Maikel P. Peppelenbosch

**Author notes:** ***Corresponding authors:*** Maikel P. Peppelenbosch, MD, PhD, Address: Dr. Molenwaterplein 40, 3015 GD, Rotterdam, The Netherlands, Zhengrong Xie, PhD, Address: The biobank, the First Affiliated Hospital of Kunming Medical University, Kunming, China, Guorong Ma, PhD, Address: School of Basic Medical, Ningxia Medical University, Yinchuan 750004, PR China.

## Abstract

**Background:** Our previous studies show that the improvement in cardiometabolic traits by intermittent fasting is associated with remodeling the gut microbiome, with short-chain fatty acids (SCFA) producing bacteria being evident. These effects, however, are largely lost when intermittent fasting is discontinued. Konjac mannan oligosaccharides (KMOS) are a commercial nature-deprived prebiotic, which has been reported to increase the levels of intestinal SCFA-producing bacteria. However, the capacity of continued KMOS consumption to maintain intermittent fasting-provoked levels of SCFA-producing bacteria, remains unknown.

**Methods:** This study aims to test whether a KMOS supplement positively affects fasting-provoked SCFA-producing bacteria levels during, and in particularly, after discontinuation of intermittent fasting. This prospective, randomized, controlled trial will be conducted in overweight volunteers aged 18-65. All participants will perform one month of intermittent fasting followed by one month of an *ad libitum* diet. Participants will be randomly assigned to receive KMOS (1.5 g/d) during fasting, both during fasting and the subsequent ad libitum period, or neither. Primary outcomes will be relative abundance of SCFA-producing bacteria in fecal samples, as determined by 16s rRNA sequencing. Secondary outcomes will be changes in body weight, blood pressure, and serum lipid levels.

**Conclusions:** Findings from this trial will answer the question whether KMOS can maintain fasting-associated SCFA producer level and metabolic benefits when fasting is discontinued.

**Clinical Trial Registration:** ChiCTR2200058139

## Introduction

Intermittent fasting (IF) has gained increasing popularity for its perceived benefits in improving weight management and enhance overall well-being in healthy individuals. An rapidly expending body of clinical research suggest demonstrate that IF also has therapeutic effects in various diseases, including diabetes (1-3), non-alcoholic fatty liver disease (4, 5), cardiometabolic disorders (6, 7), aging (8), anovulatory polycystic ovary syndrome (9), and even cancer (10-12). While the mechanism underlying these beneficial effects is not yet fully understood, remodeling of the gut microbiome is considered a potential key mechanism. Notably, IF has been shown to promote the growth of short-chain fatty acid (SCFA)-producing bacteria, particularly within the families Lachnospiraceae and Ruminococcaceae, as well as the genera *Bacteroides and Akkermansia*, which are associated with improved cardiometabolic parameters (13-20). Conversely, depletion of these bacteria has been linked to the development of the aforementioned chronic diseases (21-25), suggesting that an increase in SCFA-producing bacteria may serve as a core microbiome signature associated with the benefits of intermittent fasting.

Reducing the strictness of inclusion criteria for participants offers additional welfare benefits and facilitates broader participation in IF, allowing the procedure to be easily adopted across a diverse range of individuals within a limited timeframe. However, after cessation of IF, the beneficial effects, particularly the upregulation of SCFA-producing bacteria in the microbiome, reverts to baseline levels, which negates the observed health benefits as these microbial changes are the core mechanism with respect to these effects (15, 26, 27). Importantly, maintaining long-term IF over extended periods (e.g., over a consecutive 8-weeks) becomes increasingly difficult due to decreasing adherence and high dropout rates (28, 29). In turn, this greatly diminishes the value of IF as long-term approach to improve health. Therefore, developing strategies to sustain the key microbiome signatures associated with IF constitutes the most important open question in the field.

Konjac mannan oligosaccharides (KMOS) are commercial probiotics derived from the hydrolytic products of konjac glucomannan, originally extracted from the hemicellulose component in the cell walls of Amorphophallus konjac. KMOS primarily consists of a mixture of small-molecule polymers containing 2-20 sugar rings, polymerized by D-mannose and D-glucose through β-1,4-glycosidic bonds (30). Clinical and experimental studies suggest that KMOS supplementation can potentially improve cardiometabolic parameters (e.g., cholesterol levels, low-density lipoprotein cholesterol, and blood pressure (31-34) and colonic function and ecology (35, 36), likely due to an improvement in intestinal integrity (35). These effects are probably achieved by upregulating the expression levels of Claudin, ZO-1 and MUC-2 in intestinal epithelial cells and enhancing tight junctions (37). The upregulation of SCFA producers by KMOS (33, 35, 36, 38) may be attributed to its ability to enhance the integrity of the mucus layer, which is crucial for mucus-degrading microbiomes such as Lachnospiraceae, Ruminococcus, *Bacteroides and Akkermansia*.

Building on previous research indicating that one month of intermittent fasting (OMIF) can significantly upregulate SCFA producing bacteria in non-overweight volunteers and improve their cardiometabolic factors (15, 19), the primary aim of this trial is to evaluate whether KMOS can help sustain the upregulation of SCFA-producing bacteria and their associated beneficial effects on cardiometabolic parameters in overweight individuals after discontinuing OMIF. The secondary aim is to assess whether KMOS supplementation has an adjunctive effect on promoting SCFA producers during intermittent fasting and thereby enhancing related health benefits.

## 2. Methods

### 2.1. Study design

This randomized, open-label clinical trial aims to evaluate whether KMOS supplementation can sustain the intermittent fasting-induced proliferation of intestinal short-chain fatty acid (SCFA)-producing bacteria and the associated improvements in cardiometabolic parameters following the discontinuation of the fasting protocol.

This study was approved by the ethics committee of Northwest Minzu University in March 2022 (XBMZ-YX-20220261) and was registered at www.chictr.org.cn (ChiCTR2200058139) prior to participant’s enrollment. This study will be conducted in accordance with the principles of the Declaration of Helsinki (64th WMA General Assembly, Fortaleza, Brazil, October 2013) and the Medical Research Involving Human Subjects Act (WMO). Interested volunteers will be provided with detail information about the study and will give written informed consent prior to participation.

### 2.2. Participant recruitment

Healthy volunteers who are interested in participating in the study will be invited. To do this, flyers with contact information will be posted on social media such as WeChat and QQ. All participants expressing interest will receive a study description document that they can read and evaluate at home. Participants will be given one week to make their decision. If they have additional questions, they may consult the members of investigation team. One week later, participants will be approached again. After providing sufficient information and answering any questions, the participant who agrees to participate will be requested by the authorized person to sign the informed consent form in the presence of the coordinating investigators. The flow of the study is shown in **Figure 1**.

**Figure 1.**
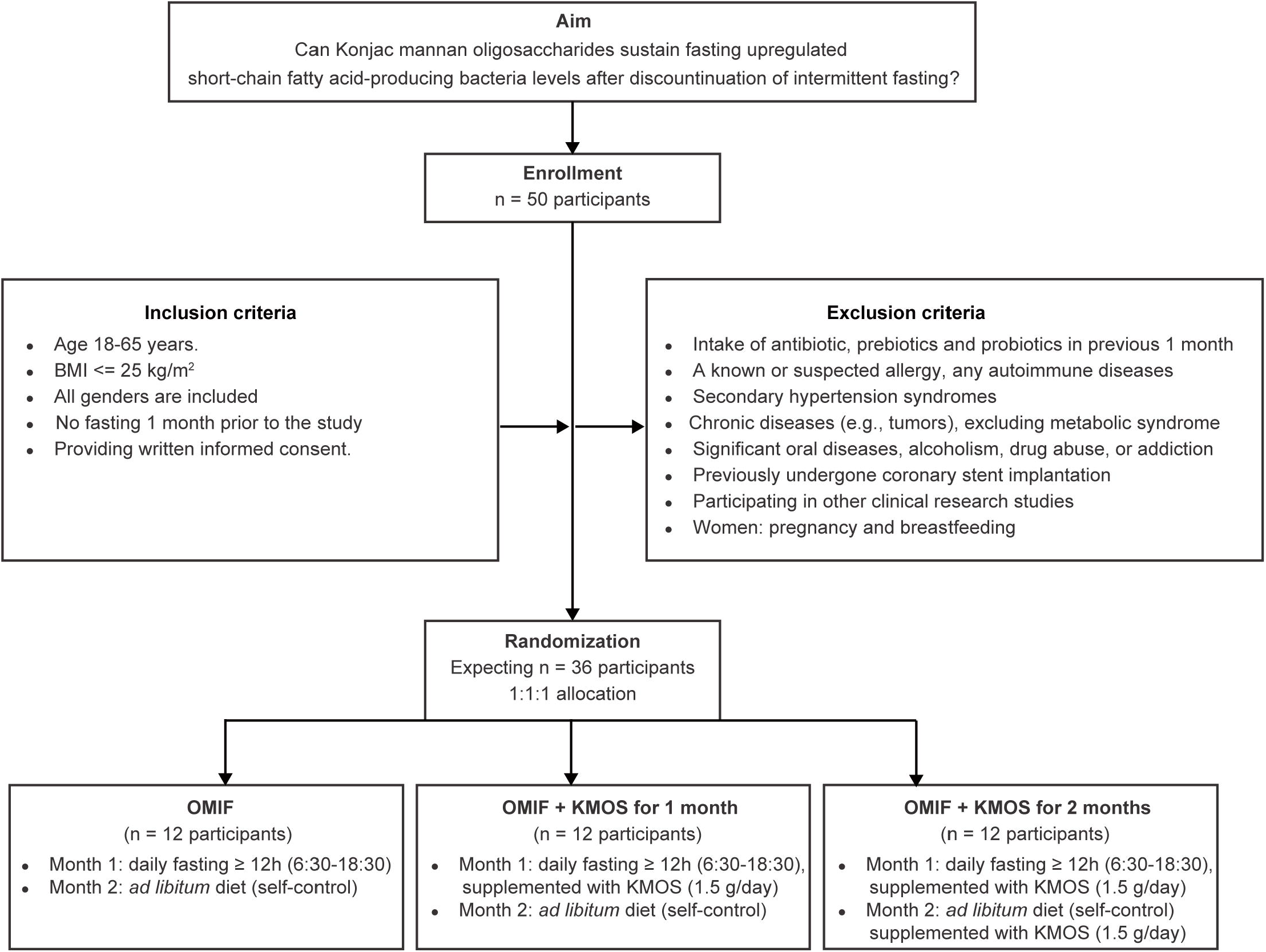
Study schema. OMIF, one month of intermittent fasting; KMOS, Konjac mannan oligosaccharides.

### 2.3. Inclusion criteria

1. Age 18-65 years
2. BMI kg/m2 > 23 (cut-off value for overweight in Asians)(39)
3. All genders will be included
4. No fasting 1 month prior to the study
5. Willing to abide by the clinical trial protocol to complete this trial, and voluntarily sign an informed consent

### 2.4. Exclusion criteria (self-report)

1. Having used antibiotics, prebiotics, or probiotics within the past month.
2. Having a known or suspected allergy.
3. Having secondary hypertension syndromes (e.g., renal parenchymal disease, renal vascular hypertension, endocrine disorders, or sleep apnea syndrome).
4. Not suffering from autoimmune diseases such as lupus erythematosus or rheumatoid arthritis.
5. Not suffering from rom chronic diseases, other as metabolic syndrome (e.g., cirrhosis, chronic nephritis, or tumors) or its sequalae.
6. Having previously undergone coronary stent implantation (cardiac stent surgery).
7. Having a current or past history of tumors, or major diseases affecting the digestive, respiratory, blood, or other systems.
8. Alcoholism, drug abuse, or addiction.
9. Having a serious nervous or mental disorder that impairs the ability to cooperate with examinations, or have suicidal tendencies.
10. Being pregnant, lactating, or planning to become pregnant soon.
11. Having significant oral diseases, such as caries, periodontal disease, or mucosal-related diseases.
12. Currently participating in other clinical research studies.
13. Any other circumstances deemed inappropriate for participation by the investigator.

### 2.5. Objective

#### 2.5.1. Primary objective

Change in the relative abundance of OMIF-associated bacteria during intermittent fasting with or without KMOS supplement (1.5 g/d). Intermittent fasting-associated bacteria mainly includes three families: Lachnospiraceae (butyrate producers) (15), Ruminococcaceae (butyrate producers) (15) and Bacteroidaceae (propionate producers) (20).

#### 2.5.2. Secondary objective(s)

Change in cardiometabolic parameters during intermittent fasting with or without KMOS supplement, in particular body composition, blood pressure, serum lipid, blood glucose ad serum insulin.

### 2.6. Power analysis

Based on our previous study on the change in the relative abundance of Lachnospiraceae in healthy volunteers underwent OMIF (15), 9 participants per group are required to have an 80% chance of detecting, as significant at the 5% level, an increase in the relative abundance of Lachnospiraceae from 0% in the control group to 15.06 % in the fasting group.

### 2.7. Study procedures (Table 1)

#### 2.7.1. Screening

During the recruitment process, a study coordinator will initially identify potential subjects via social media platforms such as WeChat and QQ. The coordinator will follow up with those who express interest and conduct a telephone screening using a screening form. Subjects who meet the initial eligibility criteria and are willing to participate will then be scheduled for a screening visit.

**Table 1.**
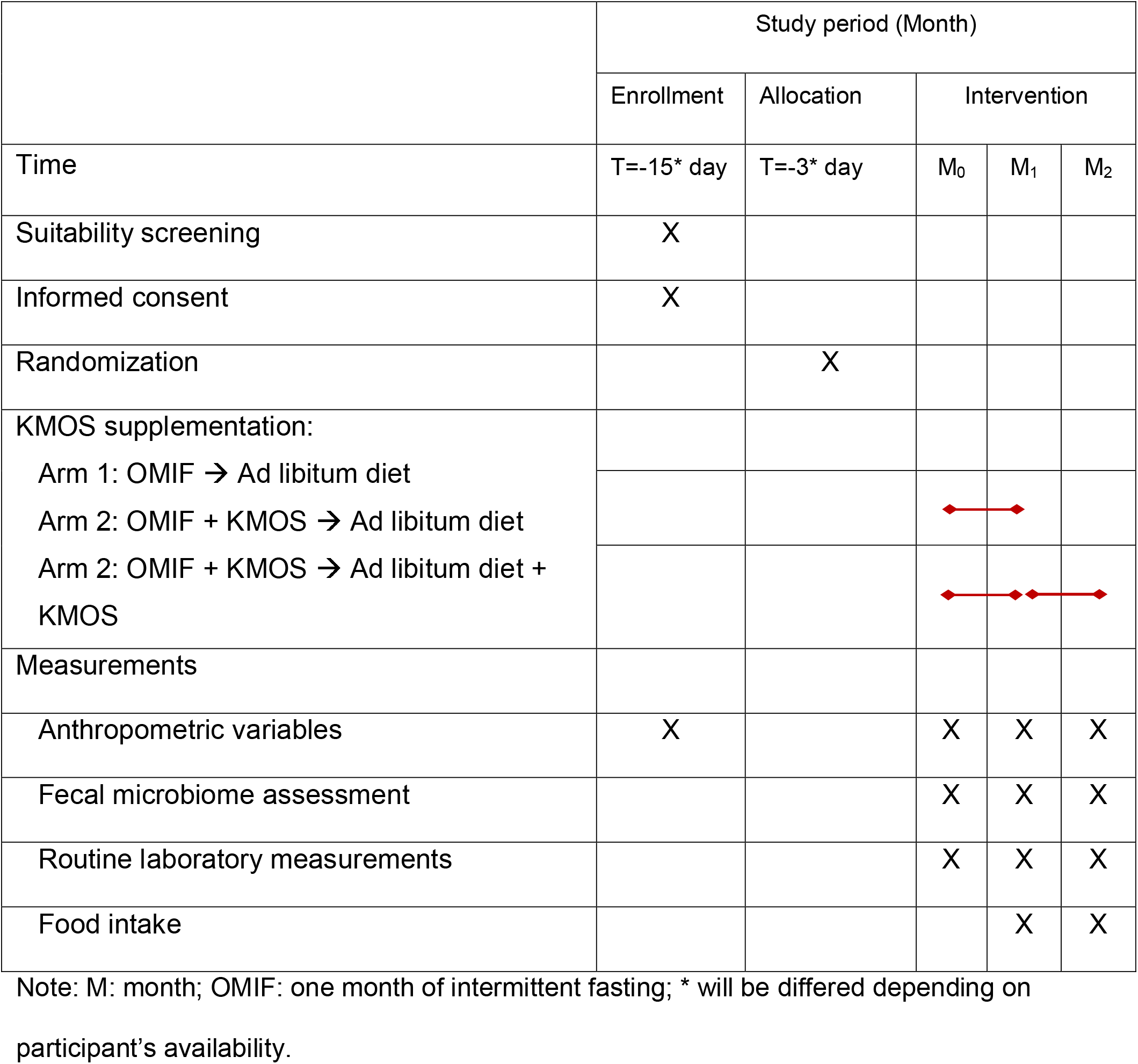
Overview of study procedures.

|  | Study period (Month) |  |  |  |  |
| --- | --- | --- | --- | --- | --- |
|  | Enrollment | Allocation | Intervention |  |  |
| Time | T=-15* day | T=-3* day | M<sub>0</sub> | M<sub>1</sub> | M<sub>2</sub> |
| Suitability screening | X |  |  |  |  |
| Informed consent | X |  |  |  |  |
| Randomization |  | X |  |  |  |
| KMOS supplementation:<br>Arm 1: OMIF → Ad libitum diet<br>Arm 2: OMIF + KMOS → Ad libitum diet<br>Arm 2: OMIF + KMOS → Ad libitum diet + KMOS |  |  |  |  |  |
| Measurements |  |  |  |  |  |
| Anthropometric variables | X |  | X | X | X |
| Fecal microbiome assessment |  |  | X | X | X |
| Routine laboratory measurements |  |  | X | X | X |
| Food intake |  |  |  | X | X |

Note: M: month; OMIF: one month of intermittent fasting; \* will be differed depending on participant's availability.

#### 2.7.2. Baseline visit

Two weeks after the screening visit, participants will be invited to visit the hospital (or alternatively a home visit by the investigating team if necessary to reach the desired inclusion) for the baseline visit to collect information on medical history, body composition (weight and BMI), blood pressure, and a fasting blood draw. Participants will be asked to provide fecal samples and information about their monthly food intake **(Table 2**).

**Table 2.** Primary and secondary outcomes.

| Outcomes | Brief description | Time frame |
| --- | --- | --- |
| Primary |  |  |
| IF-associated bacteria | 16s rRNA sequencing | One month |
| Secondary |  |  |
| BMI | Weight in kilograms and height in meters will be combined to report BMI in kg/m<sup>2</sup>. | One month |
| Blood pressure | Systolic and diastolic blood pressure will be assessed with a blood pressure monitor. | One month |
| Glucose and insulin | Blood glucose [mmol/L] and insulin [U/L] will be measured using fasting blood sample. | One month |
| Lipid profiles | Total cholesterol, HDL, LDL, Triglycerides, C-reactive protein [mmol/L] will be measured using fasting blood sample. | One month |
| Exploratory |  |  |
| Liver enzymes | Alanine transaminase, aspartate aminotransferase, gamma-glutamyl transferase and alkaline phosphatase [units/L] will be measured using fasting blood sample. | One month |
| Kidney function | Circulating concentration of creatinine [units/L] will be measured using fasting blood sample. | One month |
| Food intake | Changes in monthly calorie intake (kcal) will be converted from each food type recorded to compare the difference after OMIF versus ad libitum diets. | One month |

#### 2.7.3. Fasting intervention visit

After one month of intermittent fasting, participants will return to the hospital to provide the same information and samples as collected during baseline visit. After the completion of sample collection, participants will stop observing OMIF and return to an *ad libitum* diet.

#### 2.7.4. Post-fasting visit

One month after the cessation of intermittent fasting, participants will visit the hospital again to provide the same information and samples as collected during the baseline visit. There is no follow-up visit.

### 2.8. Microbiome analysis pipeline

#### 2.8.1. DNA extraction and 16S rRNA sequencing

##### 1) DNA extraction

Fecal DNA was extracted from the stool sample using a modified cetyltrimethylammonium bromide (CTAB) method (40). Briefly, 1000 μl of lysis buffer (CTAB) at 65°C with lysozyme will be added to a fecal sample to lyse the material. After centrifugation at 12 000 rpm for 10 min, the supernatant will be transferred into a 2.0 ml tube containing phenol/chloroform/isoamyl alcohol (25:24:1). After the centrifugation, the supernatant will be carefully transferred into a new tube. Then, 24:1 isoamyl alcohol will be added. Following a second centrifugation, the supernatant will be transferred to another tube and mixed with isopropanol. This mixture will be incubated at −20°C, after which the supernatant will be discarded. The DNA precipitate will be washed twice with 1,000 μl of 75% ethanol, then dried and dissolved in double-distilled water. Finally, 1 μl of RNase A was added, and the mixture will be incubated for 15 minutes at 37°C.

##### 2) Quality control

DNA degradation will be monitored on 1% agarose gels, and concentration will be measured using Qubit® dsDNA Assay Kit in Qubit® 2.0 Flurometer (Life Technologies, CA, USA). Samples with OD value between 1.8∼2.0 and contents above 1ug will be used for library construction.

##### 3) Library construction

The V3V4 region of the bacterial 16S rRNA gene will be amplified. PCR amplification will be carried out in 30 μl of a reaction mixture. Thermal cycling will be performed with an initial denaturation step of 1 min at 98°C followed by 30 cycles of 98°C for 10 s, 50°C for 30 s, and 72°C for 30 s, and then a final extension at 72°C for 5 min. A DNA library for next-generation sequencing will be prepared using an Ion Plus Fragment Library Kit 48 rxns (Thermo Scientific, Waltham, MA, USA) following the manufacturer’s instruction. The quality of the library will be assessed using the Qubit@ 2.0 Fluorometer (Thermo Scientific). Finally, the library will be sequenced on an Illumina platform.

#### 2.8.2. 16S rRNA raw data processing

Single end reads (250 bp) will be assigned to each sample using unique barcodes, after which the barcode and primer sequence will be removed. Quality filtering of the raw reads will be conducted under specific filtering conditions (41) to obtain high-quality clean reads. The reads will be then compared to a reference database using the UCHIME algorithm (42) to remove chimera sequences (43). Sequence analyses will be performed with the Uparse software (42). Sequences with ≥97% similarity will be clustered into the same operational taxonomy unit (OUT). Representative sequences, for each OUT will be screened for further annotation. A rarefied feature table will be generated at one depth of sequence per sample, and all downstream analyses will be performed using this rarified OTU table.

#### 2.8.3. Microbiome characterization

##### 1) Analysis of the microbiome diversity

Gut microbiota diversity will be estimated by calculating the Shannon and Simpson indices using alpha-diversity.py in QIIME (15), and the Wilcoxon signed-rank test will be used to determine statistical significance between groups. Beta diversity will be analyzed using UniFrac, which is a metric distance used for comparing microbial communities in fecal material (44). Briefly, we use weighted UniFrac (quantitative) accounts for estimating the relative abundance of each taxon within a microbial community, whereas unweighted UniFrac (qualitative) focuses on the presence/absence of taxa (45). Principal Coordinates Analysis (PCoA) of unweighted UniFrac distances will be used to indicate the corresponding dissimilarities in community membership for establishing the shift of the gut microbiota composition after intermittent fasting with or without KSOP, which will be performed using the “vegan” package in R programming language (version 4.0.2). Multivariate data analysis according to analysis of similarities (ANOSIM) will be applied to determine gut microbial shifts. A P value less than 0.05 will be considered statistically significant.

##### 2) Microbiome taxonomic analysis

To identify bacterial taxa whose sequences will be differentially abundant between groups, linear discriminant analysis (LDA) coupled with effect size measurements (LEfSe) analysis will be applied with the significance level of 0.05 and the logarithmic LDA score threshold greater than 2. Differences in OTU number and taxonomic relative abundance will be further confirmed by the Wilcoxon signed-rank test using the function wilcox.test() in R. Spearman correlation between gut microbiota and host cardiometabolic markers will be calculated using the function cor.test() in R. A P value of less than 0.05 is considered statistically significant.

### 2.9. Blood pressure measurement

Participants will be asked to sit quietly with their backs supported, without crossing their legs, and with both arms supported at heart level for 5 min before the measurements will be done. Blood pressure (BP) will be measured by an automated BP monitor with two readings at 1 min intervals, giving a total of two readings at each visit.

### 2.9. Statistical analysis

The change of relative abundance of intermittent fasting associated bacteria will be the primary objective of this study, which will be compared using repeated measurement ANOVA tests to determine whether there are statistically significant differences among three paired samples. In this paired sample set, the same participants are measured three times at three different time points. Secondary study parameters will include the measurement of cardiometabolic parameters. Repeated measurement ANOVA tests will also be used to evaluate whether there are statistically significant differences in these parameters among three dependent samples. P values <= 0.05 will be considered a statistically significant rejection of the zero hypothesis.

## 3.0. Discussion

Although implementing IF poses fewer restrictions on participant inclusion criteria, maintaining full adherence to the IF protocol over an extended or lifelong period remains a significant challenge for all participants. Notably, when IF is discontinued, the beneficial effects it provokes are largely lost (15, 19). Consequently, new strategies that could help sustain these effects are urgently needed. We hypothesize that supplementing with prebiotics may help preserve the IF-associated benefits, particularly the increased levels of intestinal SCFA-producing bacteria. KMOS will be used for this purpose, as it has been extensively studied for its prebiotic properties, including its ability to upregulate SCFA producers and improve overall well-being in humans (33, 36).

Currently, there are limited clinical trials investigating the adjunctive effects of KMOS supplementation on maintaining upregulated SCFA-producing bacteria following intermittent fasting, as well as on various cardiometabolic parameters such as body composition, blood pressure, serum lipid profiles, and glucose levels in overweight individuals. This trial has the potential to identify an easy-to-implement dietary approach to sustain the benefits of IF in this population. If successful, the findings could inform future clinical study designs, lead to revisions of dietary guidelines aimed at maintaining IF-induced upregulation of SCFA-producing bacteria, and ultimately promote overall health and well-being in participants.

## Data Availability

All data produced in the present study are available upon reasonable request to the authors

## Acknowledgement

This study is funded in part through a grant (KICH2.V4P.22.015) of the Dutch Organization for Scientific Research and the Dutch Cancer Foundation.

## Notes

### Competing Interest Statement

The authors have declared no competing interest.

### Clinical Trial

ChiCTR2200058139

### Author Declarations

Ethics committee of Northwest Minzu University gave ethical approval for this work

